# INCREASE IN INVASIVE GROUP A STREPTOCOCCAL INFECTIONS IN CHILDREN IN THE NETHERLANDS, A SURVEY AMONG 7 HOSPITALS IN 2022

**DOI:** 10.1101/2022.12.13.22283399

**Authors:** Evelien B. van Kempen, Patricia C.J. Bruijning-Verhagen, Dorine Borensztajn, Clementien L. Vermont, Marjolijn S.W. Quaak, Jo-Anne Janson, Ianthe Maat, Kim Stol, Bart J.M. Vlaminckx, Jantien W. Wieringa, Nina M. van Sorge, Navin P. Boeddha, Mirjam van Veen

**Affiliations:** Department of Paediatrics, Juliana Children’s Hospital, Haga Hospital, The Hague, The Netherlands; Department of Paediatrics, University Medical Center Utrecht, Utrecht, the Netherlands; Department of Paediatrics, Maasstad Ziekenhuis, Rotterdam, the Netherlands; Department of Paediatrics, Noordwest Ziekenhuisgroep, Alkmaar & Den Helder, the Netherlands; Department of Paediatrics, Division of Paediatric Infectious Diseases & Immunology, Erasmus MC-Sophia Children′s Hospital, University Medical Center Rotterdam, Rotterdam, The Netherlands; Department of Paediatrics, Maastricht University Medical Center+, Maastricht, the Netherlands; Department of Microbiology, Radboud University Medical Center, Nijmegen, the Netherlands; Department of Paediatrics, Division of Paediatric Infectious Diseases & Immunology, Amalia Children’s Hospital, Radboud University Medical Center, Nijmegen, the Netherlands; Department of Microbiology, St. Antonius Hospital, Nieuwegein, the Netherlands; Department of Paediatrics, Haaglanden Medical Center, The Hague, The Netherlands; Department of Medical Microbiology and Infection Prevention, Amsterdam UMC location University of Amsterdam, Amsterdam, the Netherlands; Netherlands Reference Center for Bacterial Meningitis, Amsterdam UMC location AMC, Amsterdam, the Netherlands

**Keywords:** *Streptococcus pyogenes*, invasive group A *Streptococcus*, child, outcome, epidemiology

## Abstract

Based on a survey sent to seven Dutch hospitals, we observed an substantial increase in invasive group A streptococcal infections in children in the Netherlands, comparing the pre-COVID-19 pandemic cohort of 2018-2019 to 2021-2022. The most affected age group were children between 0-5 years. Main diagnosis was pneumonia with empyema. Necrotizing fasciitis and streptococcal toxic shock syndrome were also reported in 11% and 7% respectively. A significant number was admitted to the Paediatric Intensive Care Unit. Vigilance is needed.

## Introduction

Since early 2022, pediatricians in the Netherlands have been alerted about a rise in pediatric cases of severe invasive group A streptococcal (iGAS) disease, including at least seven deaths (1,2). This coincided with an increase in the number of notifiable iGAS cases in national surveillance (3). However, this national surveillance only captures necrotizing fasciitis, toxic shock syndrome and puerperal fever/sepsis, whereas other clinical presentations of iGAS, such as (complicated) pneumonia or sepsis, are not systematically recorded. We performed a survey regarding pediatric iGAS cases in seven Dutch hospitals to determine whether case numbers were indeed elevated compared to pre-COVID-19 and to gain more insight into the clinical presentation and severity of these cases. Furthermore, we assessed the *emm*-type of invasive *Streptococcus pyogenes* isolates from pediatric iGAS cases that were received by the Netherlands Reference Laboratory for Bacterial Meningitis (NRLBM) for 2021-2022 as part of routine bacteriological surveillance.

## Methods

### Hospital survey

Clinicians from seven hospitals jointly set-up a survey to collect data on pediatric iGAS cases that were admitted to their hospital or that deceased upon arrival at their emergency department (ED). Cases were defined as: patients 0-18 years with *S. pyogenes* positive culture or PCR from an otherwise sterile body site, or a clinical diagnosis of toxic shock syndrome or necrotizing fasciitis supported by *S. pyogenes* detection from a sterile or non-sterile body site. Cases were included between July 1^st^ 2021 and June 30^th^ 2022, to capture recent trends, and compared to cases from a reference period defined as the most recent pre-COVID-19 years i.e. January 1^st^ 2018 until December 31^st^ 2019.

The participating hospitals were a convenience sample from a previous research network and included three university hospitals with Pediatric Intensive Care Unit (PICU) and four regional hospitals. The geographical location of the participating hospitals, as well as total number of hospitals with a PICU in the Netherlands is depicted in SDC figure 1.

**Figure 1:**
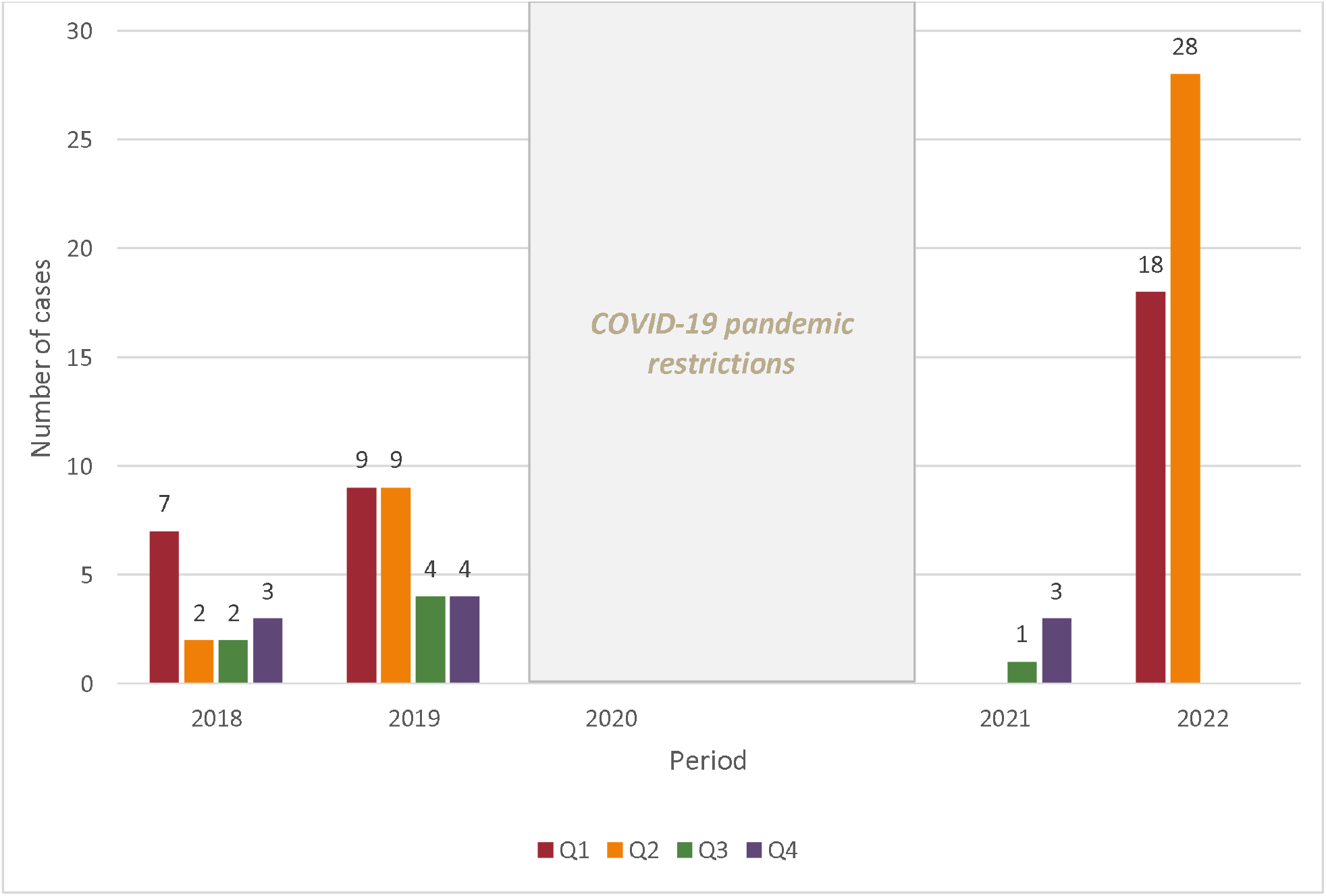
Quarterly number of pediatrics cases of invasive group A streptococcal infections (iGAS) in 2018-2019 and between July 2021 and June 2022. *Legend*: Quarterly number of paediatric invasive group A streptococcal infections (iGAS) in 2018-2019 and between July 2021 and June 2022. The in-between period was characterized by COVID-19 restrictions and not taken into account. Cases per time period (quarter) for both periods were available from 5/7 hospitals

The survey collected anonymous, aggregated data on the total number of iGAS cases per age-category (<5; 5-9 and ≥ 10 years) and per year-quarter, clinical diagnosis on admission and outcome (mortality and/or PICU admission). For cases that occurred in 2021/2022, data on coinciding or preceding (within 14 days) PCR-confirmed influenza and anamnestic evidence of recent varicella infection were also collected.

### Emm-typing of iGAS isolates

To monitor the distribution of circulating *emm*-types, the NRLBM performs *emm*-typing of invasive *S. pyogenes* isolates since 2019. The NRLBM routinely receives isolates from 1) notifiable cases of iGAS, 2) from other non-notifiable clinical iGAS presentations from nine sentinel hospitals covering ∼28% of the population, and 3) any hospital interested in typing invasive *S. pyogenes* isolates. The number and *emm*-(sub)types of *S. pyogenes* isolates received from hospitals participating in the survey and obtained from pediatric iGAS cases (not at patient level) were assessed. As surveillance started in 2019, no *emm*-types of iGAS isolates were available for the 2018-2019 reference period.

## Results

Between 1^st^ of July 2021 and 30^th^ of June 2022 (i.e. 12 months), a total of 61 pediatric iGAS cases were reported (5.1/month) versus 56 cases between Jan 2018 – Dec 2019 (i.e. 24 months, 2.3/month). The highest numbers were reported during Q1 and Q2 of 2022. Age distribution and clinical presentation were available for six out of seven hospitals. Period of presentation by annual quarter and outcome were available for six hospitals in the year 2021/2022 and five in 2018-2019. Data on preceding varicella or influenza infection was reported by five hospitals. Number of cases by year-quarter are presented in Figure 1.

The majority of cases occurred in children 0-5 years old in both periods (70% in 2021-2022 vs 50% in 2018-2019, SDC table 1). Comparing the number of cases per age-group by period, we observed the biggest increase in cases in children < 5 years of age (21 vs 38 cases in 2018-2019 and the epidemiological year 2021/2022, respectively).

The dominant clinical presentation of iGAS was sepsis (n=14, 26%) in 2018-2019 and pneumonia with empyema in 2021/2022 (n=16, 28%). In 2018-2019, no cases of necrotizing fasciitis were reported. This clinical presentation represented however 11% (n=6) of diagnoses in 2021/2022.

With regard to outcome, PICU admission occurred in in 8/39 cases (21%) in 2018-2019 and in 18/57 (32%) in 2021/2022. One death was reported in 2018-2019 (1/39; 3%) versus 5/57 (9%) in 2021/2022.

A preceding varicella infection was documented in 16 out of 49 cases (33%) and involved necrotizing fasciitis (n=6), sepsis (n=2) septic arthritis (n=2), pneumonia with empyema (n=1), erysipelas (n=3), other (n=1) and not specified (n=1). Preceding influenza infection was documented in 9 out of 49 cases (18%) and involved pneumonia with empyema (n=4), STSS (n=1), sepsis (n=1), other (n=1) and not specified (n=2).

### Emm-types

In 2021/2022, the NRLBM received 32 isolates from children from the participating institutes. 12/32 (38%) isolates were *emm* 12.0, 8/32 (25%) *emm* 1.0, and 4/32 (13%) *emm* 4.0, indicating that this increase is likely not explained by a single highly-virulent *emm*-type (SDC Fig 2).

## Discussion

This survey among seven hospitals in The Netherlands suggests that the total number of pediatric iGAS infections in 2021/2022 was increased compared to the reference pre-COVID-19 period (2018-2019). In particular, a sharp increase was observed starting early 2022 with a peak of 28 cases in the 2^nd^ quartile of 2022. These numbers reflect a 3-fold and 14-fold increase in comparison to the same quarters in 2019 and 2018, respectively. The rise in pediatric iGAS cases is most pronounced in the age group 0 – 5 years. Moreover, there seems to be a shift in most common diagnoses, with pneumonia with empyema as most frequent diagnosis in 2021/2022 and six cases of pediatric necrotizing fasciitis, a clinical condition not observed in the pre-COVID-19 reference period. Mortality was 9% among cases in 2021/2022, occurring in both young and older children, which seems higher than the reported 2-3% in children with iGAS infection from high-income countries (4-6).

The underlying causes for this iGAS surge in children are not yet fully understood. Recently, the UK Health Security Agency (UKHSA) also reported an increase in iGAS disease in 2022, particularly in children under 10, and issued an alert on Dec 2^nd^ 2022 to warn parents and clinicians (7). In the 1-4 years age group the iGAS rate was 2.3/100,000 population compared to 0.5/100,000 in the pre-pandemic period (7). A hypothesis is that mitigation measures during the COVID-19 pandemic have reduced exposure to *S. pyogenes* leading to a decreased development of protective immunity in children and an increased susceptibility to (severe) infection (8). Indeed, the UKSHA reported a concurrent rise in cases of scarlet fever. In the Netherlands however, data from sentinel surveillance in Dutch primary care do not suggest this scarlet fever trend (9). Another contributing mechanism may be that post-pandemic increased activity of other viral infections in combination with heightened activity of *S. pyogenes* predispose to secondary bacterial infections. In our survey, influenza infection was confirmed in 18% of iGAS cases and varicella infection, a known risk factor for iGAS, preceded all cases of necrotizing fasciitis (4,6). In the Netherlands, varicella vaccination is not part of the national immunization program and the 2022 varicella epidemic in the Netherlands was exceptionally high (10). A third hypothesis is the evolution of more pathogenic *emm*-types or new iGAS clones. We were unable to compare the current circulating *emm*-types to pre-COVID-19 cases because surveillance only started since 2019. Similar to our findings, UKHSA reported *emm* 12 (39%) and *emm* 1 (35%) as being dominant in pediatric iGAS cases in 2022. The current findings do not suggest one particular *emm*-type is responsible for the increase in case numbers.

The survey as study design has some limitations. Limited patient data could be collected. Therefore, we are not able to characterize the full epidemiology and clinical spectrum of the current pediatric iGAS wave. Although our survey used a convenience sample of hospitals, we feel the observed increase in iGAS is representative of the national situation, as we used the same hospitals as historical controls and our findings are in line with an increase in notifiable pediatric iGAS cases on a national level (3). This observation of iGAS surge in children in the Netherlands has implications for parents and clinicians and warrants increased vigilance for invasive disease. Early recognition and prompt initiation of therapy for children with iGAS infection may be life-saving. Future studies should focus on characterizing early predictive signs and symptoms and patient risk factors that could predict a severe disease course. Moreover, further investigation should clarify whether the distribution of *S. pyogenes* strains has changed to more easily transmissible and/or more invasive *emm*-types and whether the increase in iGAS disease is also observed in more European countries.

## Conclusion

Based on a rapid hospital-based survey, there are signs of a substantial surge in severe pediatric iGAS cases in the Netherlands since early 2022, some with fatal outcome, that requires further evaluation. Clinicians and parents should be vigilant and aware of unusual pediatric presentations such as necrotizing fasciitis. Our findings urgently warrant future studies to investigate the full impact of iGAS in Europe post-COVID.

## Data Availability

The dataset used and analysed supporting the conclusions of this manuscript are available from the corresponding author on reasonable request.

## Statements

### Ethical statement

This study was conducted in accordance with the Declaration of Helsinki and Good Clinical Practice guidelines. This study was conducted in accordance with the Declaration of Helsinki and Good Clinical Practice guidelines. The ethical review board (MEC-U, reference number W18.201) deemed that the Medical Research Involving Human Subjects Act (WMO) does not apply to our study.

### Author Approval

all authors have seen and approved the manuscript

### Funding statement

not applicable

## Acknowledgements

The authors would like to thank the members of the Dutch Study Group on Invasive Group A Streptococcal Infections in Children for their ongoing support and work.

## Conflict of interest

**NMvS** declares royalties related to a patent (WO 2013/020090 A3) on vaccine development against *Streptococcus pyogenes* (Vaxcyte; Licensee: University of California San Diego with **NMvS** as co-inventor). All other authors report no conflicts of interest.

## Authors’ contributions

MvV, NPB and EBvK conceptualised and designed the study. EBvK analysed the data. PCJBV, DB, BJMV, JWW, NMvS, MvV, NPB, EvK wrote the manuscript. CLV, MSWQ, JJ, IM, KS, DB, BJMV, JWW, NMvS, MvV, NPB, EvK collected the data. All authors critically reviewed the manuscript and approved it for publication. MvV and NPB supervised the conduct of the study.

## Figure legend

**Supplemental digital content Figure 1:**
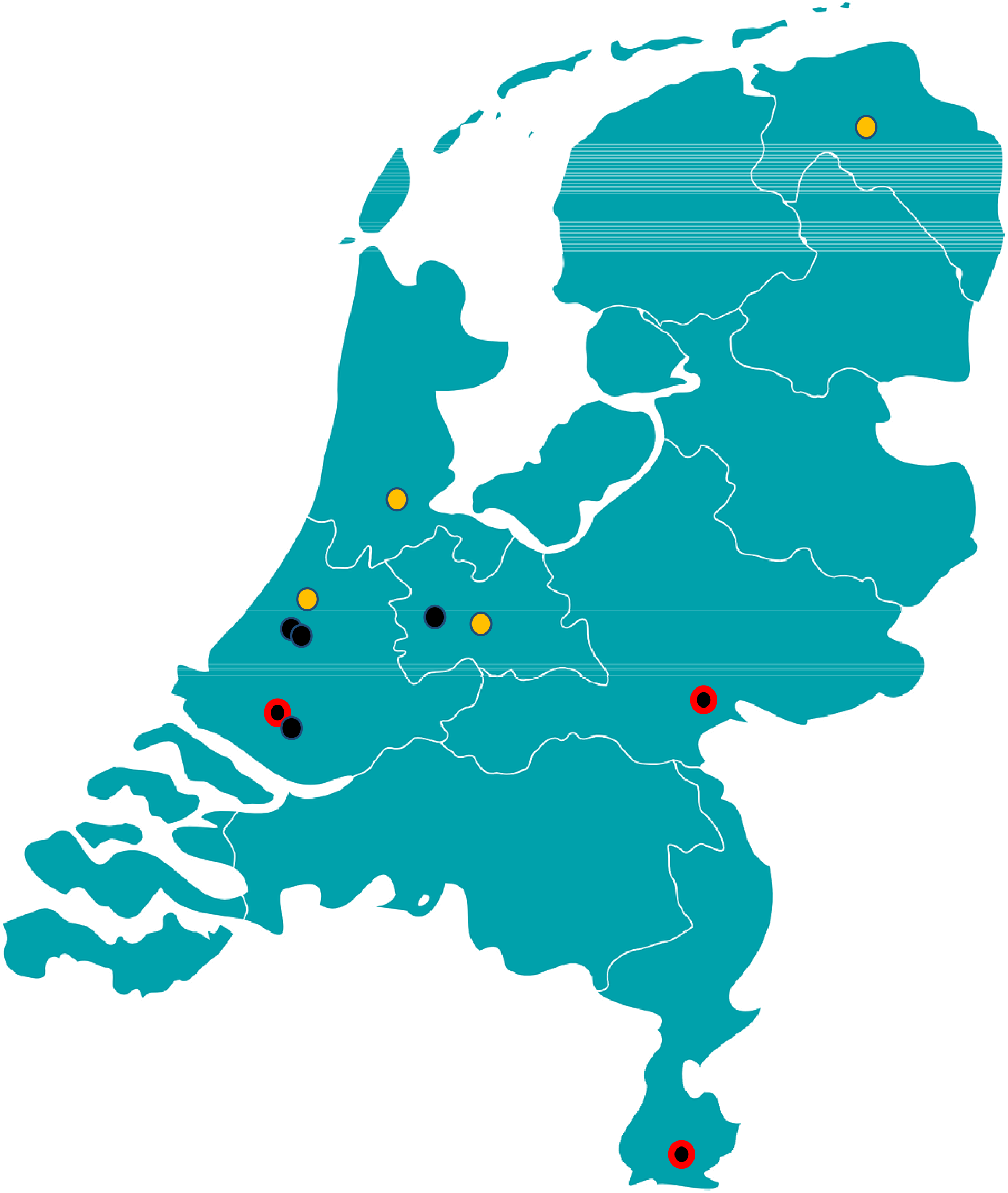
Geographical location of the participating hospitals, as well as total number of hospitals with a Pediatric Intensive Care Unit. 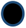 Indicate location of participating regional hospitals 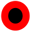 Indicate location of participating hospitals with Pediatric Intensive Care Unit 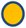 Indicate location of hospital with Pediatric Intensive Care Unit

**Supplemental Digital Content (SDC) - Table 1:**
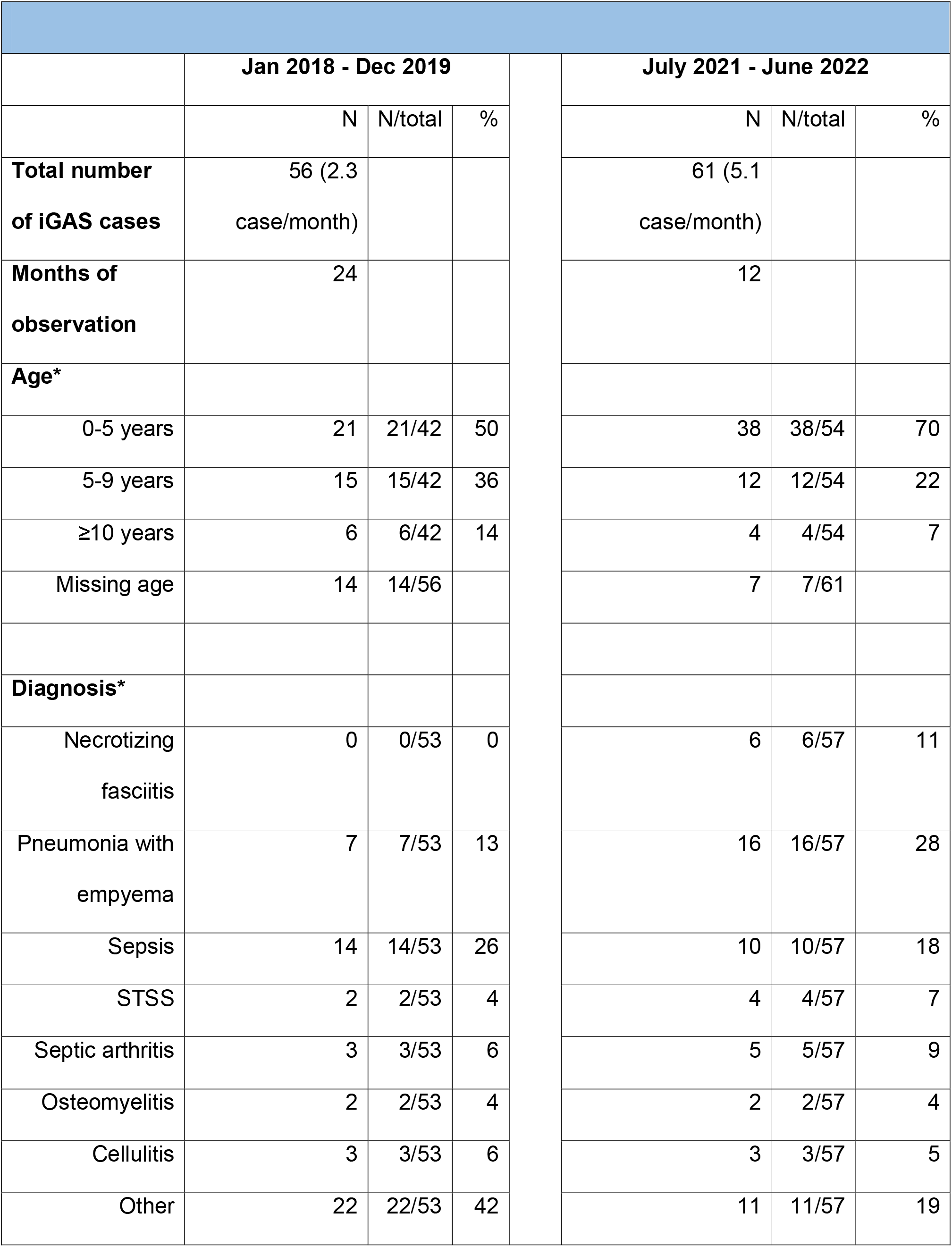
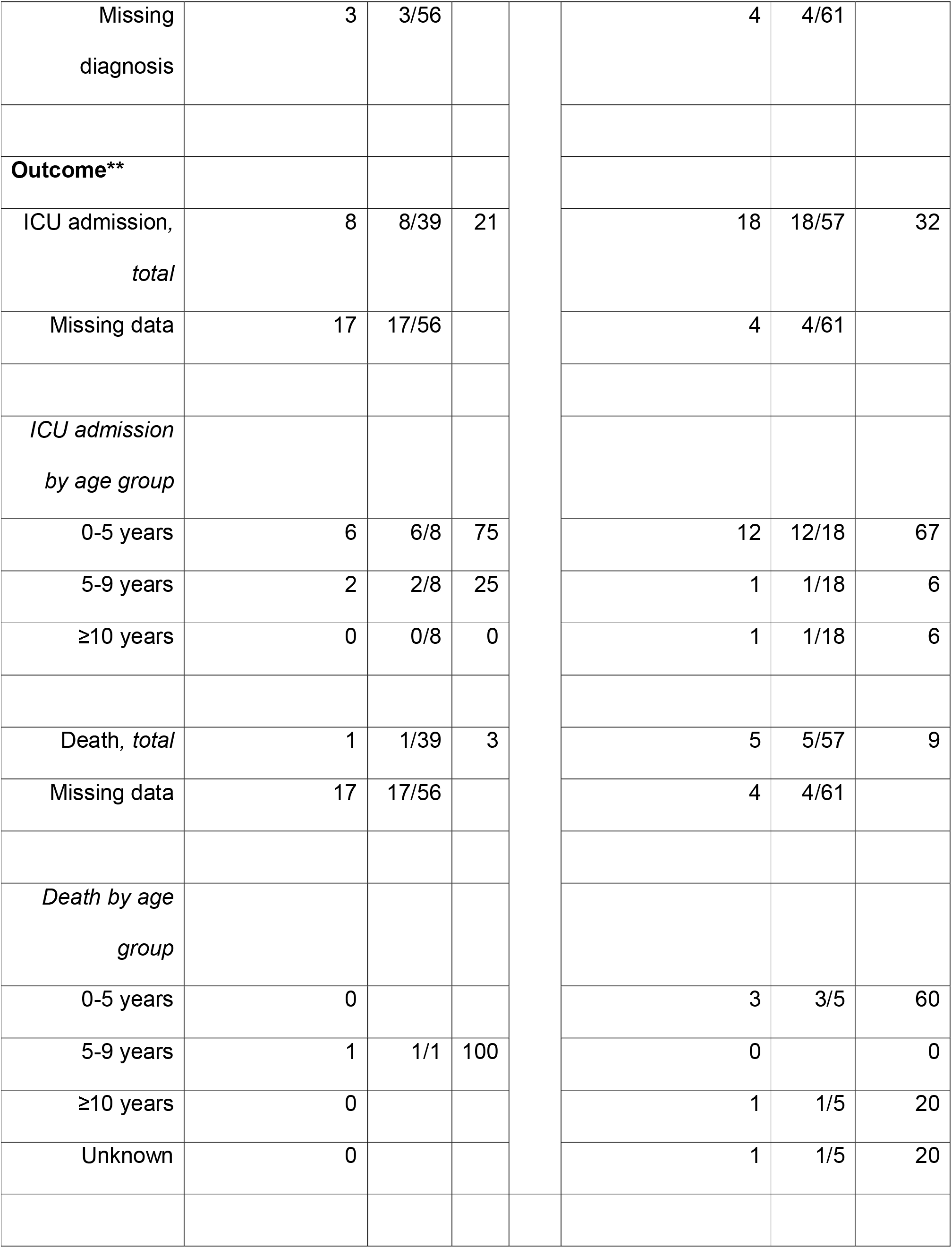
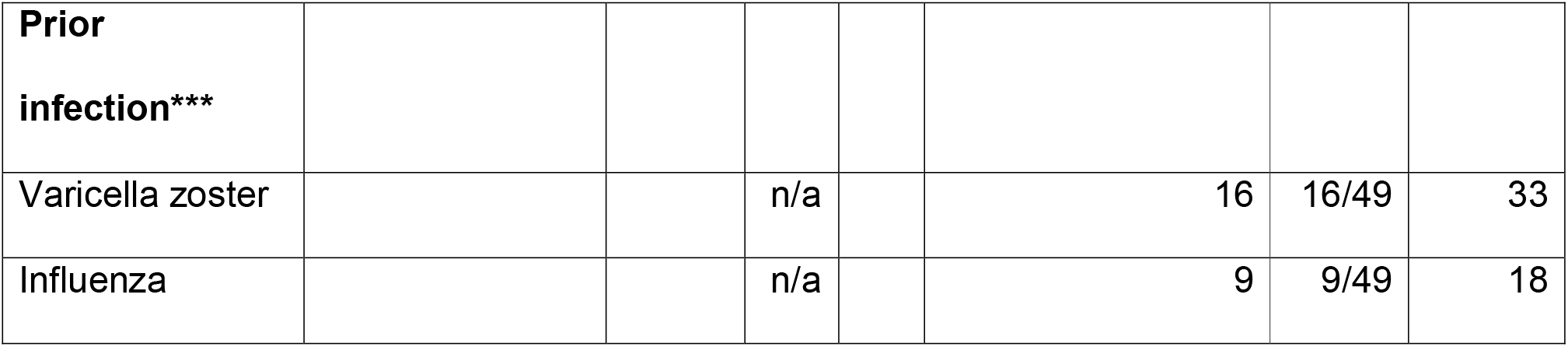
Digital Content (SDC) - Characteristics of 117 pediatric cases with invasive group A streptococcal infections (iGAS) in the two study periods: Jan 2018-Dec 2019 (i.e. 24 months) and July 2021-June 2022 (i.e. 12 months). **Table 1:** Age, clinical presentation, and outcome of invasive Group A streptococcal infections in the pediatric cohorts 2018-2019 and 2021-2022. * Data available from 6/7 hospitals ** Data available from 5/7 hospitals in 2018-2019 and 6/7 hospitals in 2021-2022 *** Data available from 5/7 hospitals Other diagnoses for both periods include: tonsillitis with bacteremia, mastoiditis with bacteremia, cervical lymphadenitis, abscesses at different locations, bacterial tracheitis, meningitis and peritonitis.

**Supplemental Digital Content (SDC) - Figure 2:**
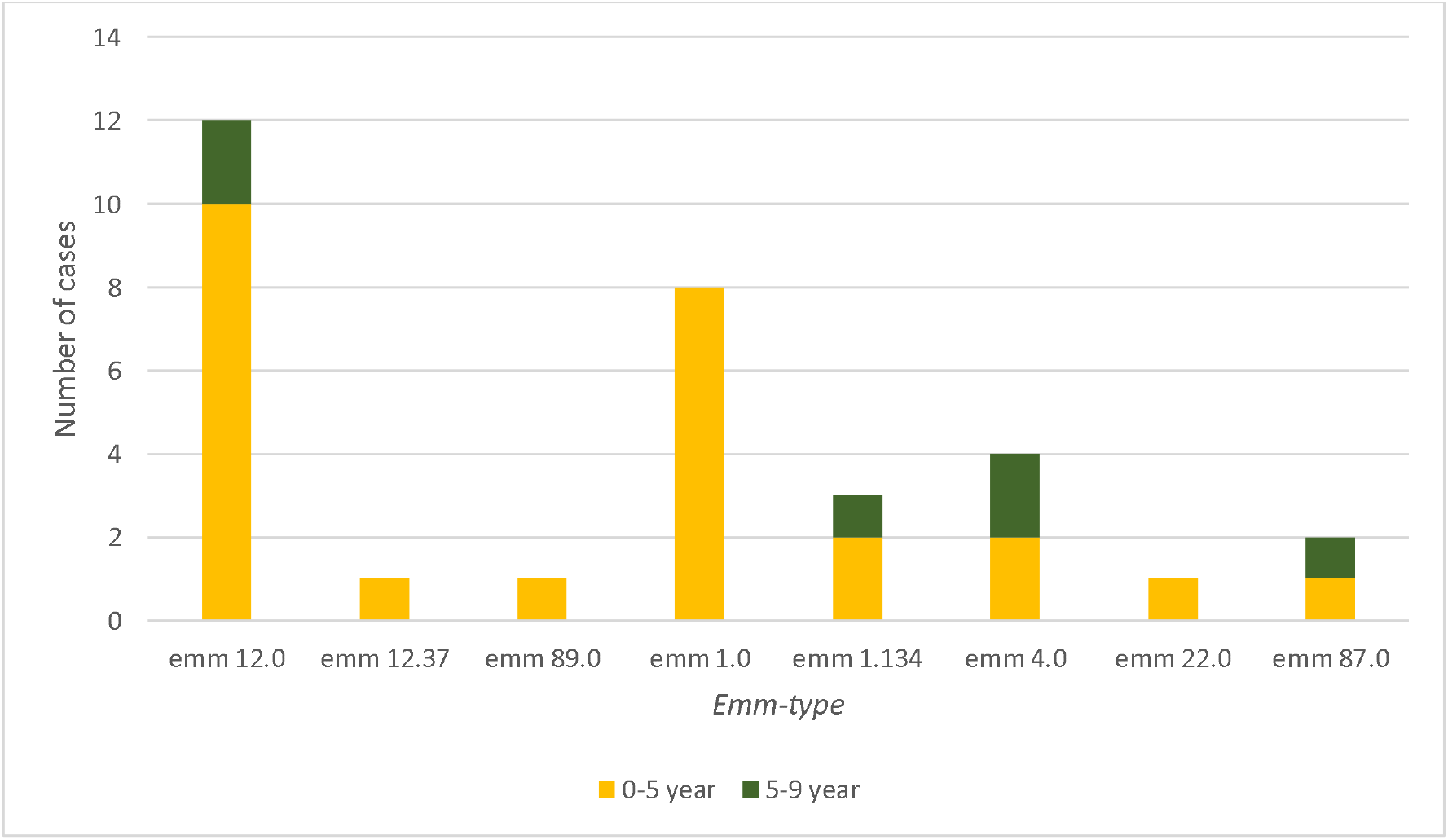
Digital Content (SDC) - Distribution of *emm*-types of 32 *S. pyogenes* isolates received by the National Reference Laboratory for Bacterial Meningitis in July 2021 – June 2022. Legend: In 2021-2022, the National Reference Laboratory Bacterial Meningitis received 32 isolates from children 0-18 years from the participating institutes.

## Notes

### Funding Statement

This study did not receive any funding

### Author Declarations

Ethics committee of Medical research Ethics Committees United the Netherlands (MEC-U) gave ethical approval for this work This study was conducted in accordance with the Declaration of Helsinki and Good Clinical Practice guidelines. This study was conducted in accordance with the Declaration of Helsinki and Good Clinical Practice guidelines. The ethical review board (MEC-U, reference number W18.201) deemed that the Medical Research Involving Human Subjects Act (WMO) does not apply to our study

